# Evaluation of whole blood point-of-care placental growth factor testing for prediction of adverse maternal and perinatal outcomes: a prospective observational cohort study in Sierra Leone

**DOI:** 10.1101/2025.03.14.25324010

**Authors:** Katy Kuhrt, Rossetta Cole, Moses M’Bayoh, Chileshe Mabula-Bwalya, Alice Hurrell, Alexandra Ridout, Cristina Fernandez-Turienzo, Paul T. Seed, Lucy C. Chappell, Kate Bramham, Andrew H. Shennan

## Abstract

**Objectives:** Pre-eclampsia is a major cause of maternal death. Placental growth factor (PlGF) testing improves time-to-diagnosis and outcomes. We evaluated two novel, whole blood, point-of-care (POC) PlGF tests (RONIA^TM^ and Lepzi® Quanti PlGF) in a low-resource setting, for prediction of adverse outcomes.

**Study Design:** A prospective observational cohort study in hypertensive pregnant women, 24–36^+6^ weeks’ gestation, at a tertiary maternity hospital in Sierra Leone.

**Methods:** Eligible, consented women underwent RONIA^TM^ and/or Lepzi® Quanti PLGF testing; results were concealed. Optimal rule-out and rule-in thresholds were determined for prediction of predefined maternal (maternal death, eclampsia) and perinatal (stillbirth, termination pre-viability, neonatal death before discharge) composite outcomes. Sensitivity, specificity, negative (NPV) and positive predictive values were determined.

**Results:** Analysis was performed on women with complete outcomes: RONIA^TM^ n=488 and Lepzi® Quanti PlGF n=140. Optimal thresholds were <60pg/mL or <90pg/mL (rule-out) and <20pg/mL or <12pg/mL (rule-in) for RONIA^TM^ and Lepzi® Quanti PlGF respectively. For tests performed <34 weeks’ gestation, RONIA^TM^ PlGF <60pg/mL had high sensitivity, 94.9% (95%CI 82.7-99.4%) and NPV, 94.6% (95%CI 81.8-99.3%) for maternal outcomes, with sensitivity, 100% (95%CI 95.8-100.0%) and NPV, 100% (95%CI 90.5-100%) for the perinatal composite. Lepzi® Quanti PlGF < 90pg/mL had 100% sensitivity and NPV for all predefined maternal and neonatal outcomes. Performance reduced slightly at later gestations.

**Conclusions:** Whole blood POC-PlGF measurement demonstrates accurate rule-out performance of two novel devices for serious outcomes, with potential for individualised risk stratification in low-resource settings.

## Introduction

Pre-eclampsia and other hypertensive disorders of pregnancy affect 10% of pregnancies(1), and are a leading cause of maternal death. It is estimated that around 40,000 maternal deaths and 500,000(1) neonatal deaths annually are associated with pre-eclampsia, the majority occurring in low-and middle-income settings (LMICs). Most deaths can be avoided with early identification, enabling timely management (antihypertensives, anticonvulsants, and appropriately planned birth).

Pre-eclampsia is often asymptomatic, even in the presence of severe disease(2), making diagnosis challenging, particularly in LMICs, due to limited access to blood pressure (BP) monitoring equipment, urine dipsticks, blood tests and ultrasound scans to assess fetal wellbeing. Even when diagnostics are available, BP and urine have poor sensitivity for predicting adverse pregnancy outcomes (18-36%) (3), limiting their clinical utility to stratify risk and determine management.

The advent of angiogenic biomarker testing, such as Placental Growth Factor (PlGF) alone and sFLT-1/PlGF ratio, has transformed the ability to identify high-risk women(4, 5), and PLGF-based testing is now incorporated into National Institute for Health and Care Excellence (NICE) national UK guidelines(6) for management of women with suspected pre-eclampsia and into International Society for the Study of Hypertension in Pregnancy (ISSHP) guidelines in the definition of pre-eclampsia(7).

The few PlGF studies that have been undertaken in LMICs, include a blinded prospective cohort study(8) and an operational pilot study of plasma PlGF measurement(9) in women presenting with suspected pre-eclampsia in Mozambique, where low PlGF concentration was significantly associated with shorter time-to-delivery and adverse pregnancy outcomes, and a small pilot study in India where 50 hypertensive women underwent third trimester SfLT-1/PlGF and high risk ratios were associated with more severe pre-eclampsia, and pregnancy complications(10).

However, to date PlGF-based measurements for routine clinical use and study settings, including in LMICs, are not suitable for point-of-care (POC), and require centrifugation. This is prohibitive to sustainable implementation in Sierra Leone, and other LMICs. RONIA^TM^ is a small, POC PlGF platform, utilizing innovative upconverting nanoparticle technology, which enables high-sensitivity, quantitative immunoassays from a single drop of whole blood (20µL), with a result in 30 minutes. Lepzi® Quanti PlGF is a small, portable, fluorescence immunoassay for quantitative determination of PlGF in EDTA anticoagulated whole blood or plasma samples, requiring 200 µL of venous whole blood, with a result in 15 minutes. Our objective was to evaluate test performance of novel whole blood RONIA^TM^ and Lepzi® Quanti PlGF devices for predicting adverse pregnancy outcomes in a low-resource setting.

## Methods

### Study design

This prospective observational cohort study was undertaken between June 2022 and April 2024 in a tertiary government maternity hospital in Freetown, Sierra Leone. Consecutive women ≥16 years old presenting or referred with symptoms or signs of suspected or actual pre-eclampsia between 24^+0^ to 36^+6^ weeks of gestation, with a singleton or twin pregnancy, were eligible. Participants were included when the attending healthcare provider deemed that the women required evaluation for pre-eclampsia, with symptoms or signs such as headache, visual disturbances, epigastric or right upper quadrant pain, hypertension, dipstick protein or suspected fetal growth restriction. Written study information was provided, explained verbally, and participants signed or marked the consent form by thumbprint for those unable to write. BP measurement was taken as part of routine care, with women sitting, and with the right arm supported at the level of the heart, using a pregnancy-validated device. BP measurement was repeated if hypertension was detected on the first reading and the second reading recorded. Normotensive readings were not repeated.

For the RONIA^TM^ test, 20 microlitres (µL) of whole blood were collected by finger-prick, and for the Lepzi® Quanti PlGF test, 200 µL of whole blood were collected by venous draw, according to manufacturers’ instructions. Both samples were immediately analysed using RONIA^TM^ and Lepzi® Quanti readers respectively to determine PlGF concentration. Results were concealed to the clinical team. Pregnancy outcome details for the woman and infant were obtained from case note review, and data quality checks were carried out by an external researcher, including adjudication by an obstetrician or neonatologist. The study was approved by Kings College London Research Ethics Committee (RECSM22/23-22669) and Sierra Leone Ethics and Scientific Review Committee (10/03/2022).

The pre-specified primary maternal outcome was a composite of maternal death or eclampsia. Additional maternal outcomes were: individual components of the composite; parameters defined in the miniPIERS consensus, where available and reported in this setting (excluding blood transfusion as this was commonly performed for reasons other than pre-eclampsia-related) (11): early delivery (<37 weeks’ gestation, within 7 and 14 days of test; severe hypertension (BP ≥160/110 mmHg); high dependency unit (HDU) admission; use of magnesium sulfate; use of intravenous antihypertensives. Perinatal outcomes included a composite of one or more of stillbirth; neonatal death before discharge, or termination pre-viability (to protect maternal life). Additional perinatal outcomes were: components of the composite; mode of delivery; gestational age at delivery (by best clinical estimate); birthweight; birthweight centile; neonatal unit admission; sepsis with clinical evidence of infection; antibiotics for possible serious bacterial infection; Apgar score at 5 and 10 min; clinical diagnosis of hypoxic ischaemic encephalopathy (low Apgar score at birth, no crying, need for resuscitation); neonatal seizures; respiratory distress syndrome; supplementary oxygen required: use of continuous positive airway pressure ventilation; clinical diagnosis of necrotising enterocolitis (abdominal distension with a history of formula feeding); hypoglycaemia (<2.6 mmol) requiring intervention; hypothermia (single documented temperature <36.5 °C); neonatal jaundice requiring phototherapy; nasogastric feeding.

### Sample size

The required sample size was calculated for accurate estimation of the sensitivity (within 10% absolute) and specificity (within 6%) of RONIA^TM^ PlGF in determining the primary maternal endpoint. We assumed a sensitivity of 0.90, specificity 0.90 and 95% confidence intervals (2-tailed), requiring 62 cases and 144 controls, and a total sample size of 488 women, based on an observed (post hoc) prevalence of 12.7%. We anticipated 10% loss to follow-up because of challenges related to the setting and adjusted our sample size accordingly (n=536). During the study period, we had an additional opportunity to collect samples for Lepzi® Quanti PlGF, and therefore a subset of women had both PlGF tests.

### Statistical Analysis

Women were classified according to their gestation at the time of the PlGF test, <34 weeks of gestation; 34 to 36^+6^ weeks of gestation, based on best clinical estimate. Descriptive statistics were used to summarise the demographic data and POC-PlGF results were presented using mean (standard deviation), or median (interquartile range) depending on distribution. Optimal rule-in and rule-out cut-points were selected from a range of thresholds taking into account the balance between sensitivity and specificity and performance for predicting maternal and perinatal composite outcomes. For these, and other pre-defined endpoints, sensitivity, specificity and positive predictive values (PPV) and negative predictive values (NPV) were calculated based on the selected rule-in and rule-out thresholds. Median and interquartile ranges from time from POC-PlGF testing to delivery were calculated. We did a sensitivity analysis on the primary maternal and perinatal outcomes excluding twin pregnancies. Analysis was performed with statistical support from King’s College London Life Course Sciences (STATA/SE Version 18). The study is reported in accordance with STAndards for the Reporting of Diagnostic accuracy studies (STARD) guidelines (Figure S1).

## Results

536 women were recruited and underwent RONIA^TM^ testing; 161 women also underwent Lepzi® Quanti PlGF testing. We recruited all those who were approached, eligible and consented (Figure 1). No adverse events were reported related to performing PlGF testing. Outcome measures were collected in full on 488/536 and 140/161 occasions for RONIA^TM^ and Lepzi® Quanti tests respectively. Women without a valid PlGF result (RONIA^TM^, n=2; LEPZI® Quanti, n=7) and those lost to follow-up (RONIA^TM^ n=46; LEPZI® Quanti, n=14) were excluded. The characteristics of the remaining women are shown in Table 1, along with maternal and perinatal outcomes, with additional adverse maternal and perinatal outcomes in supplementary tables (Tables S1 and S2 respectively). Proportions of women by hypertension category and maternal and perinatal adverse events stratified by PlGF are shown in Figure 2. The diagnostic accuracy of PlGF for predicting maternal and perinatal composite outcomes is shown in Tables 2 and 3, based on our selected thresholds, <20 and <60pg/mL for RONIA^TM^ PlGF and <12 and <90pg/mL for Lepzi® Quanti PlGF.

**Figure 1.**
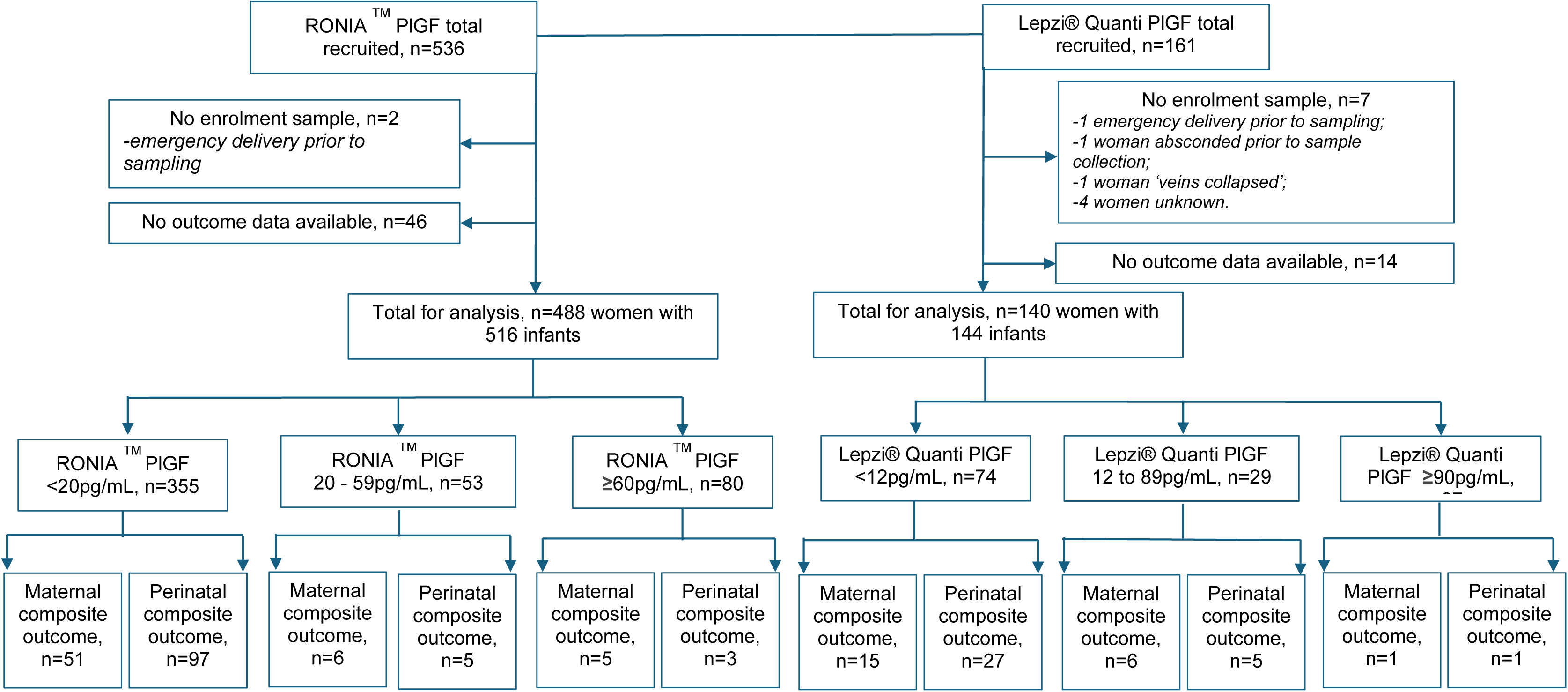
Flow diagram of participants through the study. RONIA^TM^ samples were collected from June 2022 to April 2024, Lepzi® Quanti samples were collected from July 2023 to April 2024. Therefore 140 women have both RONIA^TM^ and Lepzi® Quanti samples available for analysis.

**Figure 2.**
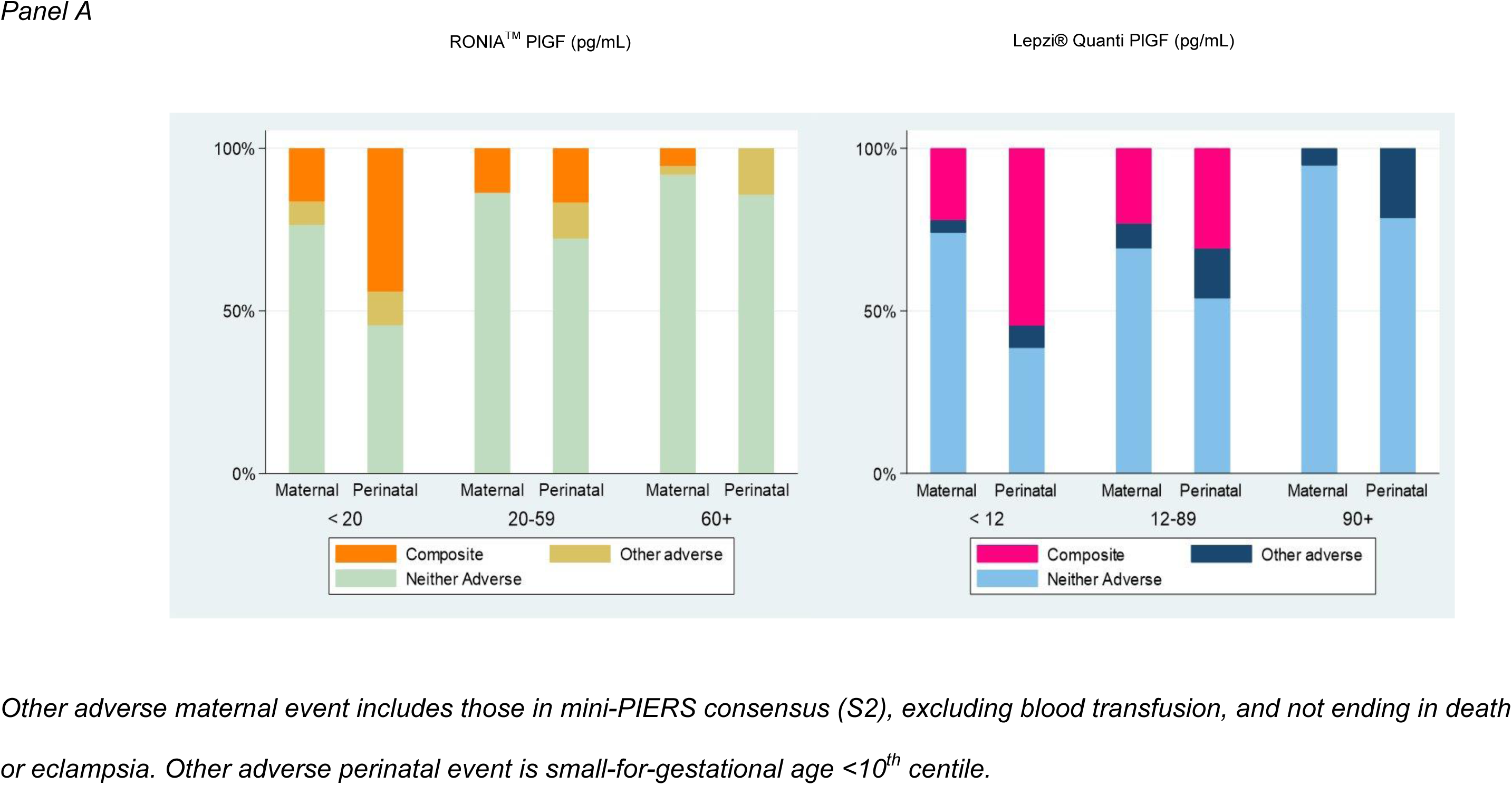
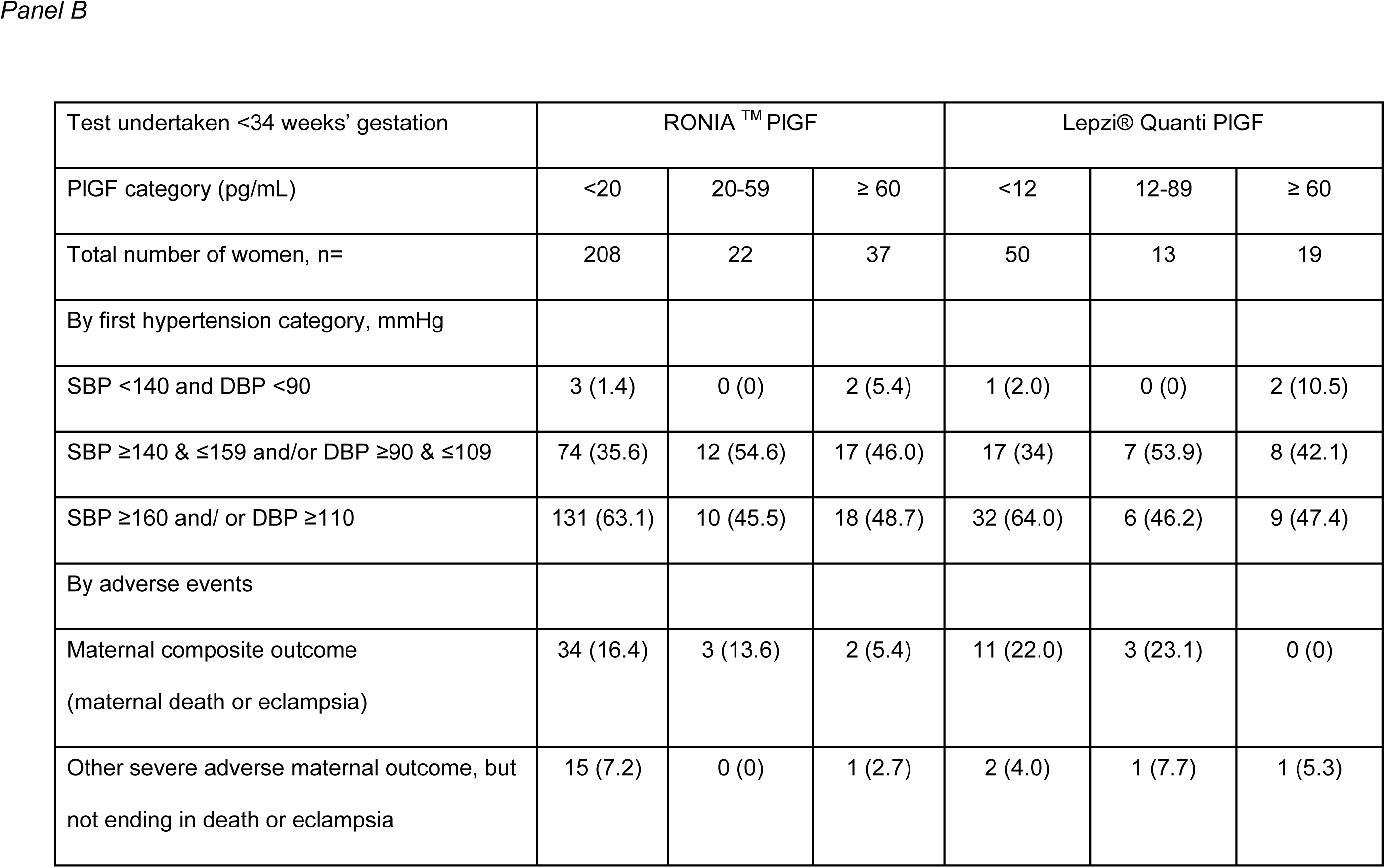
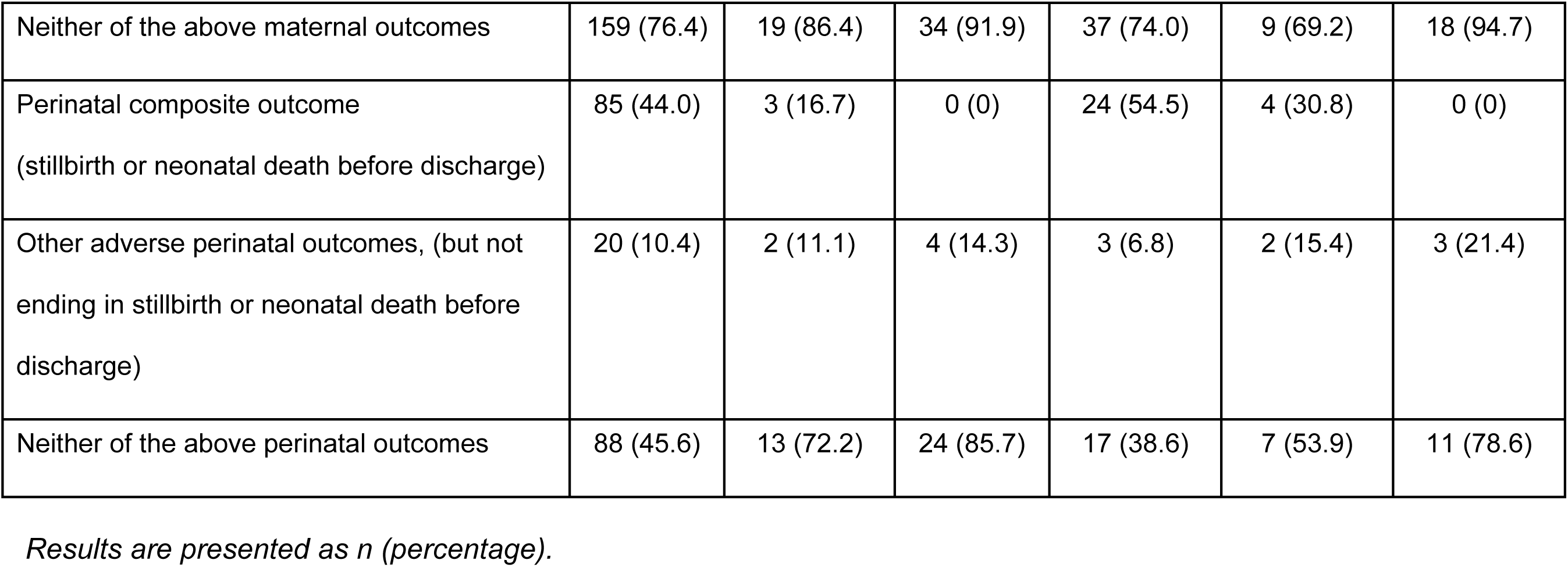
Hypertension category and adverse maternal and perinatal events stratified by PlGF category in women with tests done at <34 weeks’ gestation.

**Table 1.** Maternal demographics, admission characteristics, delivery details and maternal and perinatal outcomes.

|  | RONIA™ PIGF |  |  | Lepzi® Quanti PIGF |  |  |
| --- | --- | --- | --- | --- | --- | --- |
| Gestation at PIGF testing, weeks | <34 | 34-36 | All | <34 | 34-36 | All |
| Demographic details, n | 267 | 221 | 488 | 82 | 58 | 140 |
| Age , y | 27.0 [6.3] | 27.3 [6.2] | 27.1 [6.3] | 27.6 [5.9] | 27.2 [6.7] | 27.4 [6.2] |
| Primiparous | 92 (35.0) | 73 (33.0) | 165 (34.1) | 25 (30.5) | 25 (43.1) | 50 (35.7) |
| Admission characteristics |  |  |  |  |  |  |
| Gestation at enrolment, weeks | 32.0<br>(29.0-34.0) | 36.0<br>(35.0-36.0) | 33.0<br>(30.0-36.0) | 30.0<br>(28.0-32.0) | 36.0<br>(35.0-36.0) | 32.0<br>(29.0-36.0) |
| Admission/ first SBP, mmHg | 162 [20] | 158 [21] | 160 [21] | 163 [22] | 156 [25] | 160 [23] |
| Admission/ first DBP, mmHg | 109 [15] | 107 [16] | 108 [16] | 110 [15] | 108 [16] | 109 [16] |
| Maximum SBP, mmHg | 174 [19] | 172 [22] | 173 [21] | 176 [17] | 173 [26] | 175 [21] |
| Maximum DBP, mmHg | 119 [17] | 117 [18] | 118 [17] | 121 [18] | 117 [17] | 120 [18] |
| Admission urine dipstick, n | 203 | 172 | 375 | 53 | 33 | 86 |
| 1+ | 40 (19.7) | 27 (15.7) | 67 (17.9) | 12 (22.6) | 3 (9.1) | 15 (17.4) |
| 2+ | 34 (16.7) | 31 (18.0) | 65 (17.3) | 14 (26.4) | 6 (18.2) | 20 (23.3) |
| 3+ | 102 (50.2) | 65 (37.8) | 167 (44.5) | 21 (39.6) | 14 (42.4) | 35 (40.7) |
| Nil | 27 (13.3) | 49 (28.5) | 76 (20.3) | 6 (11.3) | 10 (30.3) | 16 (18.6) |
| Admission creatinine, n | 69 | 74 | 143 | 2 | 10 | 12 |
| Highest creatinine | 90.0<br>(71.6-111.0) | 76.0<br>(67.0-96.5) | 81.7<br>(68.3-105.0) | 70.0<br>(68.0-72.0) | 61.0<br>(54.0-67.0) | 64.5<br>(55.0-70.0) |
| Highest creatinine $\geq 77$ $\mu\text{mol/L}$ | 47 (68.1) | 36 (48.6) | 83 (58.0) | 0 (0) | 0 (0) | 0 (0) |
| Delivery details and maternal outcomes |  |  |  |  |  |  |
| Twin pregnancy | 14 | 18 | 32 | 3 | 5 | 8 |
| Gestation at delivery, weeks | 32.0 | 36.0 | 34.0 | 31.5 | 36.0 | 34.0 |
|  | (29.0-34.0) | (35.0-37.0) | (32.0-36.0) | (28.0-33.0) | (35.0-36.0) | (31.0-36.0) |
| Preterm birth <34 weeks | 183 (72.3) | 0 (0.0) | 183 (38.7) | 61 (78.2) | 0 (0.0) | 61 (45.2) |
| Preterm birth <37 weeks | 234 (92.5) | 159 (72.9) | 393 (83.4) | 69 (88.5) | 48 (84.2) | 117 (86.7) |
| Maternal outcomes, n |  |  |  |  |  |  |
| Composite maternal outcome | 39 (14.6) | 23 (10.4) | 62 (12.7) | 14 (17.1) | 8 (13.8) | 22 (15.7) |
| Maternal death | 12 (4.5) | 5 (2.3) | 17 (3.5) | 6 (7.3) | 1 (1.7) | 7 (5.0) |
| Eclampsia | 30 (11.2) | 20 (9.0) | 50 (10.2) | 10 (12.2) | 8 (13.8) | 18 (12.9) |
| Severe Hypertension (SBP ≥ 160/110 mmHg) | 225 (84.3) | 162 (73.3) | 387 (79.3) | 73 (89.0) | 46 (79.3) | 119 (85.0) |
| Mode of birth, n | 264 | 221 | 485 | 80 | 58 | 140 |
| Caesarean section | 124 (47.0) | 126 (57.0) | 250 (51.5) | 35 (43.8) | 32 (55.2) | 67 (47.9) |
| Vaginal birth | 140 (53.0) | 95 (43.0) | 235 (48.5) | 45 (56.3) | 26 (44.8) | 71 (50.7) |
| Resource dependent outcomes |  |  |  |  |  |  |
| HDU Admission* | 16 (6.1) | 7 (3.2) | 23 (4.8) | 6 (7.3) | 1 (1.8) | 7 (5.1) |
| Use of magnesium sulfate | 237 (91.0) | 179 (81.4) | 416 (86.5) | 74 (90.2) | 48 (82.8) | 122 (87.1) |
| Use of IV antihypertensives | 128 (50.6) | 75 (35.2) | 203 (43.6) | 57 (71.2) | 25 (44.6) | 82 (60.3) |
| Perinatal outcomes (where recorded), n | 277 | 239 | 516 | 82 | 63 | 145 |
| Composite perinatal outcome | 89 (32.1) | 18 (7.5) | 107 (20.7) | 28 (34.1) | 5 (7.9) | 33 (22.8) |
| Stillbirth | 74 (26.7) | 14 (5.9) | 88 (17.1) | 21 (25.6) | 4 (6.3) | 25 (17.2) |
| Termination pre-viability for maternal health | 3 (1.1) | 0 (0) | 3 (0.6) | 0 (0) | 0 (0) | 0 (0) |
| Live birth | 200 (712.2) | 225 (94.1) | 425 (82.4) | 61 (74.4) | 59 (93.7) | 120 (82.8) |
| Neonatal unit admission | 88/185 (47.6) | 42/218 (19.3) | 130/403 (32.3) | 26/59 (44.1) | 12/58 (20.7) | 38/117 (32.5) |
| Neonatal death before discharge | 12/145 (8.3) | 4/202 ( 2.0) | 16/347 (4.6) | 7/54 (13.0) | 1/56 (1.8) | 8/110 (7.3) |
| Live births, n | 200 | 225 | 425 | 61 | 59 | 120 |
| Apgar score at 1 minute | 6 (4-7) | 8 (6-8) | 7 (5-8) | 5 (3-7) | 8 (6-8) | 7 (5-8) |
| Apgar score at 5 minutes | 7 (5-8) | 9 (7-9) | 8 (6-9) | 6 (3-8) | 9 (7-9) | 8 (6-9) |
| Birthweight | 2000<br>(1500-2400) | 2800<br>(2300-3100) | 2400<br>(1900-2900) | 2000<br>(1400-2400) | 2500<br>(2200-2900) | 2300<br>(1900-2700) |
| SGA (<10<sup>th</sup> customized birthweight centile) | 31/174 (17.8) | 27/204 (13.2) | 58/378 (15.3) | 9/51 (17.6) | 9/55 (16.4) | 18/106 (17.0) |
| SGA (<3<sup>rd</sup> customized birthweight centile) | 13/174 (7.5) | 17/204 (8.3) | 30/378 (7.9) | 4/51 (7.8) | 6/55 (10.9) | 10/106 (9.4) |
| SGA (<1<sup>st</sup> customized birthweight centile) | 8/174 (4.6) | 7/204 (3.4) | 15/378 (4.0) | 4/51 (7.8) | 2/55 (3.6) | 6/106 (5.7) |

*Results are presented as mean [standard deviation]; n (percentage); median (IQR).*
*\*only four HDU, high dependency beds available at any one time. PIGF, placental growth factor.*

**Table 2.** RONIA^TM^ and Lepzi® Quanti PlGF prediction of composite maternal outcome based on PlGF thresholds of <20pg/mL and <60pg/mL, and <12pg/mL and <90pg/mL respectively.

|  | Composite maternal outcome (death or eclampsia) |  |  |  |  |  |
| --- | --- | --- | --- | --- | --- | --- |
|  | RONIA™ PIGF |  |  | Lepzi® Quanti PIGF |  |  |
| Gestation at testing, weeks | <34 | 34-36<sup>+6</sup> | All | <34 | 34-36<sup>+6</sup> | All |
| Prevalence, n (%) | 39 (14.6) | 23 (10.4) | 62 (12.7) | 14 (17.1) | 8 (13.8) | 22 (15.7) |
| PIGF threshold | <20pg/mL<br>% (95%CI) |  |  | <12pg/mL<br>% (95%CI) |  |  |
| Sensitivity | 87.2<br>(72.6-95.7) | 73.9<br>(51.6-89.8) | 82.3<br>(70.5-90.8) | 78.6<br>(49.2-95.3) | 50.0<br>(15.7-84.3) | 68.2<br>(45.1-86.1) |
| n/N | 34/39 | 17/23 | 51/62 | 11/14 | 4/8 | 15/22 |
| Specificity | 23.7<br>(18.3-29.7) | 34.3<br>(27.8-41.4) | 28.6<br>(24.4-33.2) | 42.6<br>(30.7-55.2) | 60.0<br>(45.2-73.6) | 50.0<br>(40.7-59.3) |
| n/N | 54/228 | 68/198 | 122/426 | 29/68 | 30/50 | 59/118 |
| Positive predictive value | 16.3<br>(11.6-22.1) | 11.6<br>(6.9-17.9) | 14.4<br>(10.9-18.5) | 22.0<br>(11.5-36.0) | 16.7<br>(4.7-37.4) | 20.3<br>(11.8-31.2) |
| n/N | 34/208 | 17/147 | 51/355 | 11/50 | 4/24 | 15/74 |
| Negative predictive value | 91.5<br>(81.3-97.2) | 91.9<br>(83.2-97.0) | 91.7<br>(85.7-95.8) | 90.6<br>(75.0-98.0) | 88.2<br>(72.5-96.7) | 89.4<br>(79.4-95.6) |
| n/N | 54/59 | 68/74 | 122/133 | 29/32 | 30/34 | 59/66 |
| PIGF threshold | <60pg/mL |  |  | <90pg/mL |  |  |
| Sensitivity | 94.9<br>(82.7-99.4) | 87.0<br>(66.4-97.2) | 91.9<br>(82.2-97.3) | 100.0<br>(76.8-100.0) | 87.5<br>(47.3-99.7) | 95.5<br>(77.2-99.9) |
| n/N | 37/39 | 20/23 | 57/62 | 14/14 | 7/8 | 21/22 |
| Specificity | 15.4<br>(10.9-20.7) | 20.7<br>(15.3-27.0) | 17.8<br>(14.3-21.8) | 27.9<br>(17.7-40.1) | 34.0<br>(21.2-48.8) | 30.5<br>(22.4-39.7) |
| n/N | 35/228 | 41/198 | 76/426 | 19/68 | 17/50 | 36/118 |
| Positive predictive value | 16.1<br>(11.6-21.5) | 11.3<br>(7.0-16.9) | 14.0<br>(10.8-17.8) | 22.2<br>(12.7-34.5) | 17.5<br>(7.3-32.8) | 20.4<br>(13.1-29.5) |
| n/N | 37/230 | 20/177 | 57/407 | 14/63 | 7/40 | 21/103 |
| Negative predictive value | 94.6<br>(81.8-99.3) | 93.2<br>(81.3-98.6) | 93.8<br>(86.2-98.0) | 100.0<br>(82.4-100.0) | 94.4<br>(72.7-99.9) | 97.3<br>(85.8-99.9) |
| n/N | 35/37 | 41/44 | 76/81 | 19/19 | 17/18 | 36/37 |

*Results are presented as n (percentage); percentage (95 percent confidence interval). PIGF, placental growth factor.*

**Table 3.**
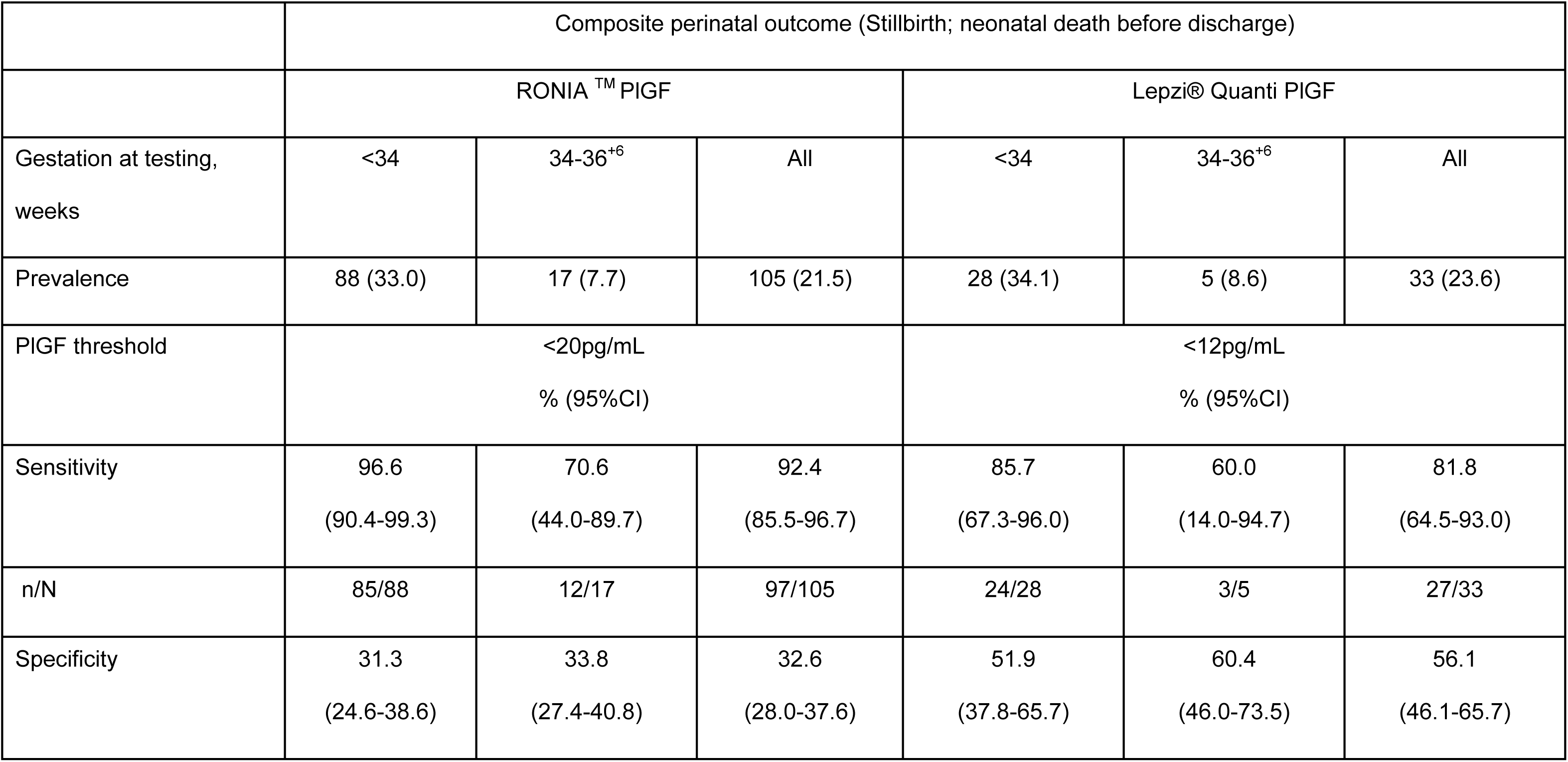
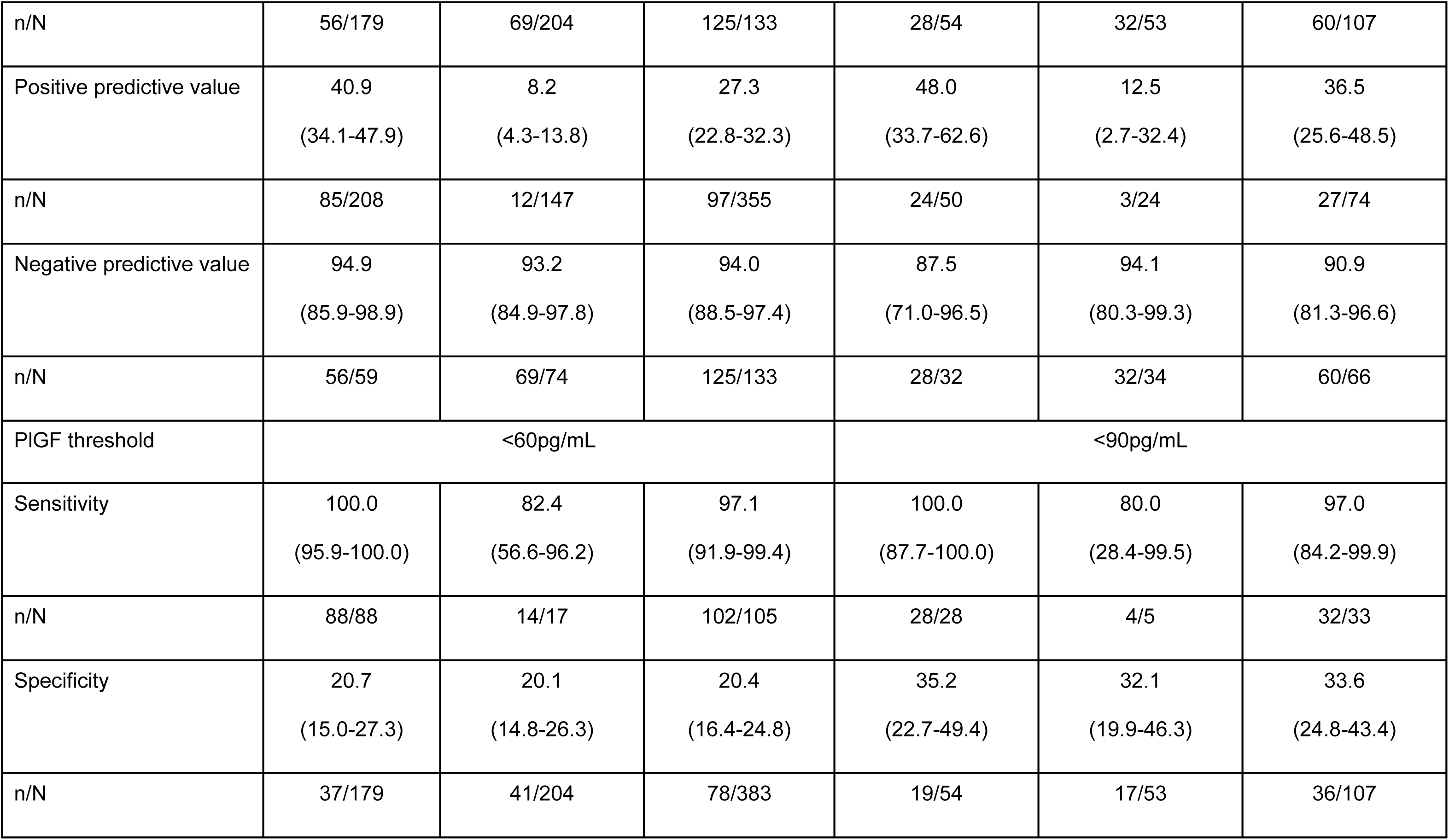
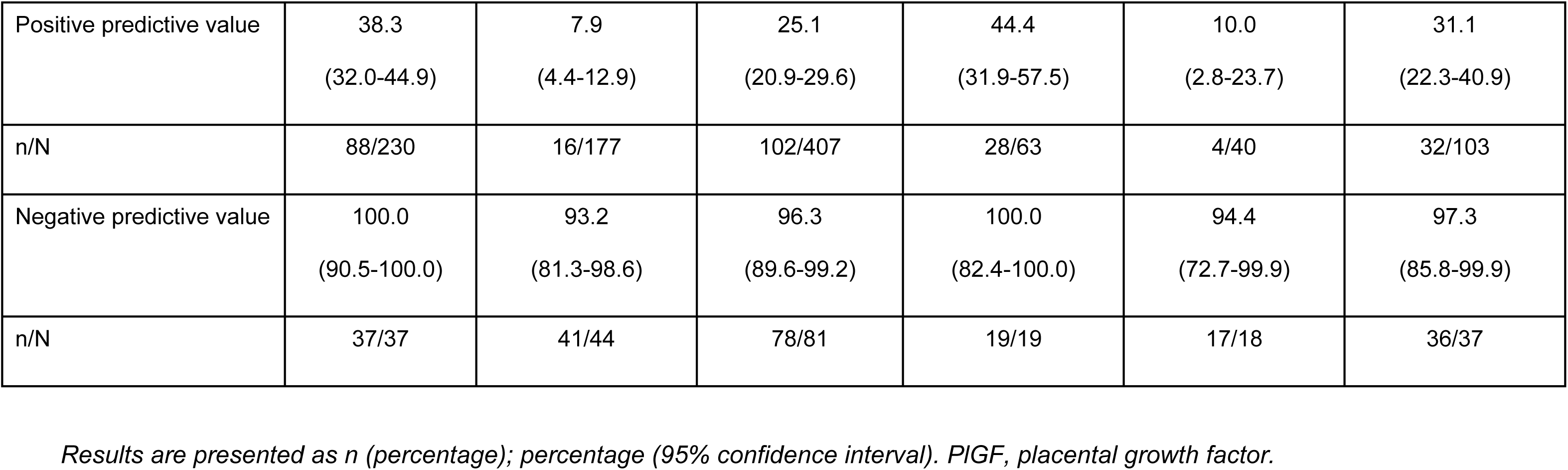
RONIA^TM^ and Lepzi® Quanti PlGF prediction of composite perinatal outcome based on PlGF thresholds of <20pg/mL and <60pg/mL, and <12 and <90pg/mL respectively.

For the maternal composite (n=62), abnormal RONIA^TM^ PlGF <60pg/mL had high sensitivity, 91.9% (95%CI 82.2-97.3%) and NPV, 93.8% (95%CI 86.2-98.0%) for women tested at all gestations. Sensitivity and NPV for the maternal composite improved still further for women tested at <34 weeks’ gestation to 94.9% (95%CI 82.7-99.4%) and 94.6% (95%CI 81.8-99.3%) respectively, and for maternal death (n=17), where sensitivity and NPV were 100%. Abnormal Lepzi® Quanti PlGF <90pg/mL had 100% sensitivity and NPV for the maternal composite (n=22) (Table 2), and for individual components of the composite, declining slightly at later gestations, except for maternal death (n=7) where 100% sensitivity and NPV were maintained across all gestations. (Tables S3 and S4).

For the perinatal composite outcome (n=105), abnormal RONIA^TM^ PlGF <60pg/mL had high sensitivity, 97.1% (95%CI 91.8-99.4%) and NPV, 96.3% (95%CI 89.6-99.2%) at all gestations tested, and these were improved in women tested <34 weeks’ gestation with sensitivity and NPV of 100%, (95%CIs 95.8-100.0% and 90.5 to 100% respectively (Table 4). Lepzi® Quanti PlGF had 100% sensitivity and NPV for the perinatal composite outcome (Table 4), and for the individual components of the composite, for women tested at <34 weeks’ gestation, declining slightly at later gestations tested, except for neonatal death before discharge, where 100% sensitivity and NPV were maintained across all gestations tested (Tables S5-6).

Amongst women who underwent RONIA^TM^ testing at all gestations, there were eight false negative cases (one maternal death; four eclampsia; one stillbirth and two neonatal deaths prior to discharge), three of which may have been associated with alternative or additional diagnoses (such as coexistent infectious disease, or a possible seizure disorder), whilst sepsis was reported in one of the babies who had a neonatal death (with normal birthweight). For Lepzi® Quanti PlGF, there were two false negative cases (one eclampsia case, PlGF: 276.44pg/mL; one severe adverse neonatal event, PlGF: 92.68pg/mL). Both of these cases were verified against the source notes, but due to limited documentation, it was not possible to determine whether additional diagnoses may have contributed to or caused their outcomes.

Prediction of additional clinical outcomes is presented in supplementary tables S7-S11. For both abnormal RONIA ^TM^ <60pg/mL and Lepzi® Quanti <90pg/mL, sensitivity and NPV was 100% for women tested <34 weeks for SGA <3^rd^ centile, albeit in small numbers of women (Table S8).

For RONIA^TM^ PlGF <20pg/mL, PPV for delivery within 14 days was 88.0% (95%CI 84.0-91.2%), and for preterm delivery <37 weeks’ gestation it was 87.1% (95%CI 83.0-90.4%), which improved to 97.4% (95%CI 94.0-99.1%) for women tested <34 weeks’ gestation. For RONIA^TM^ PlGF <60, although sensitivity was high (>90%), for both delivery within 14 days and preterm delivery <37 weeks’ gestation, for women tested <34 weeks’ gestation, NPV was modest due to high prevalence of the outcome (S10, S11). For Lepzi® Quanti PlGF <12pg/ml, PPV for prediction of delivery within 14 days was 87.2% (95%CI 74.3-95.2%) for women tested <34 weeks’ gestation, improving at later gestations. For preterm delivery <37 weeks’ PPV was 97.8% (95%CI 88.5-99.9%) for women tested <34 weeks’ gestation, declining at later gestations. However, despite modest overall performance in predicting time to delivery, median time between test and delivery was markedly longer in women with a normal PlGF result for both tests (Figure 3): RONIA^TM^ (34 vs 4 days); Lepzi® Quanti (21.5 vs 3 days). A prespecified sensitivity analysis excluding women with twin pregnancies showed very similar results (Tables S12 and S13).

**Figure 3.**
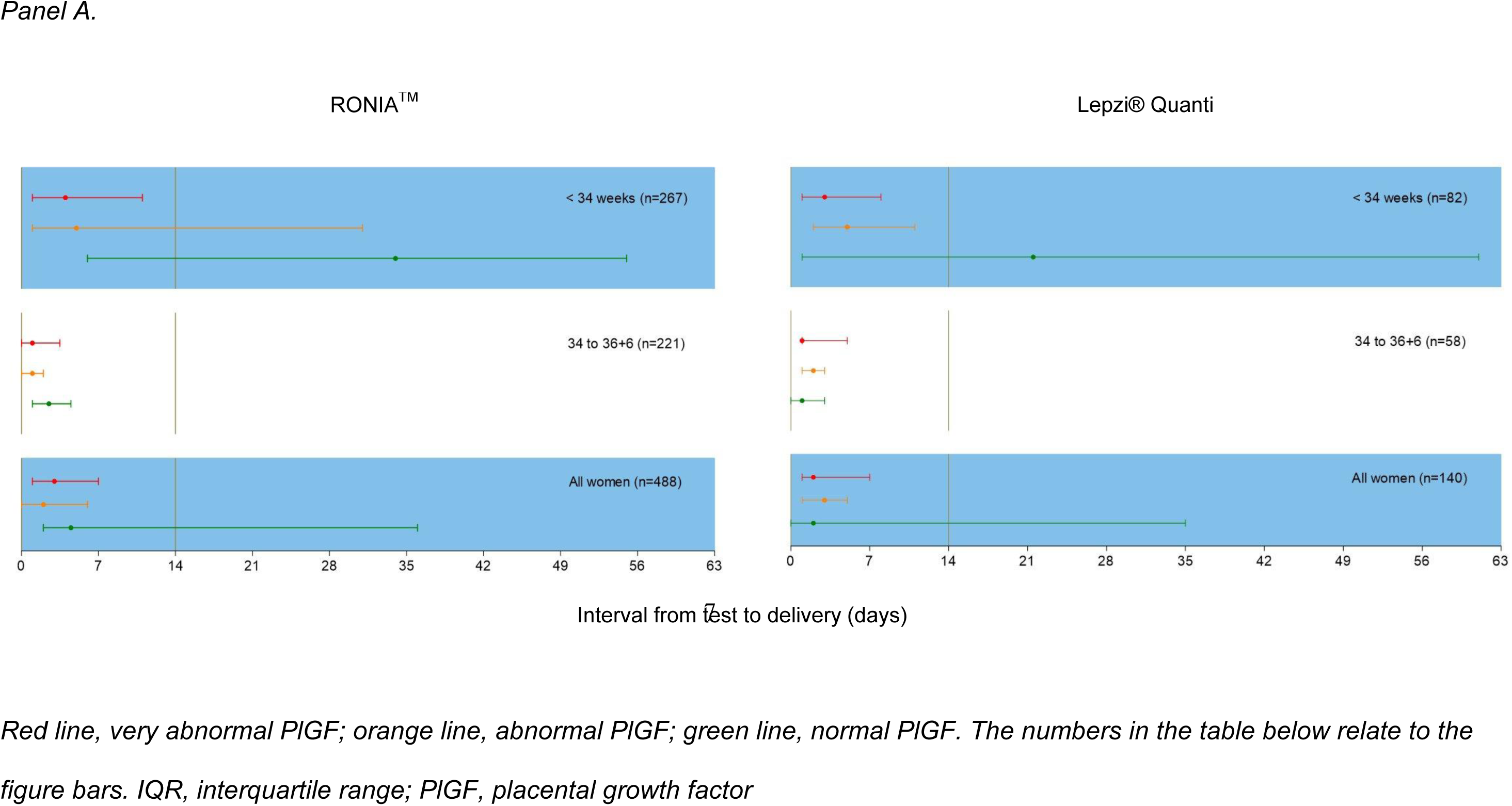
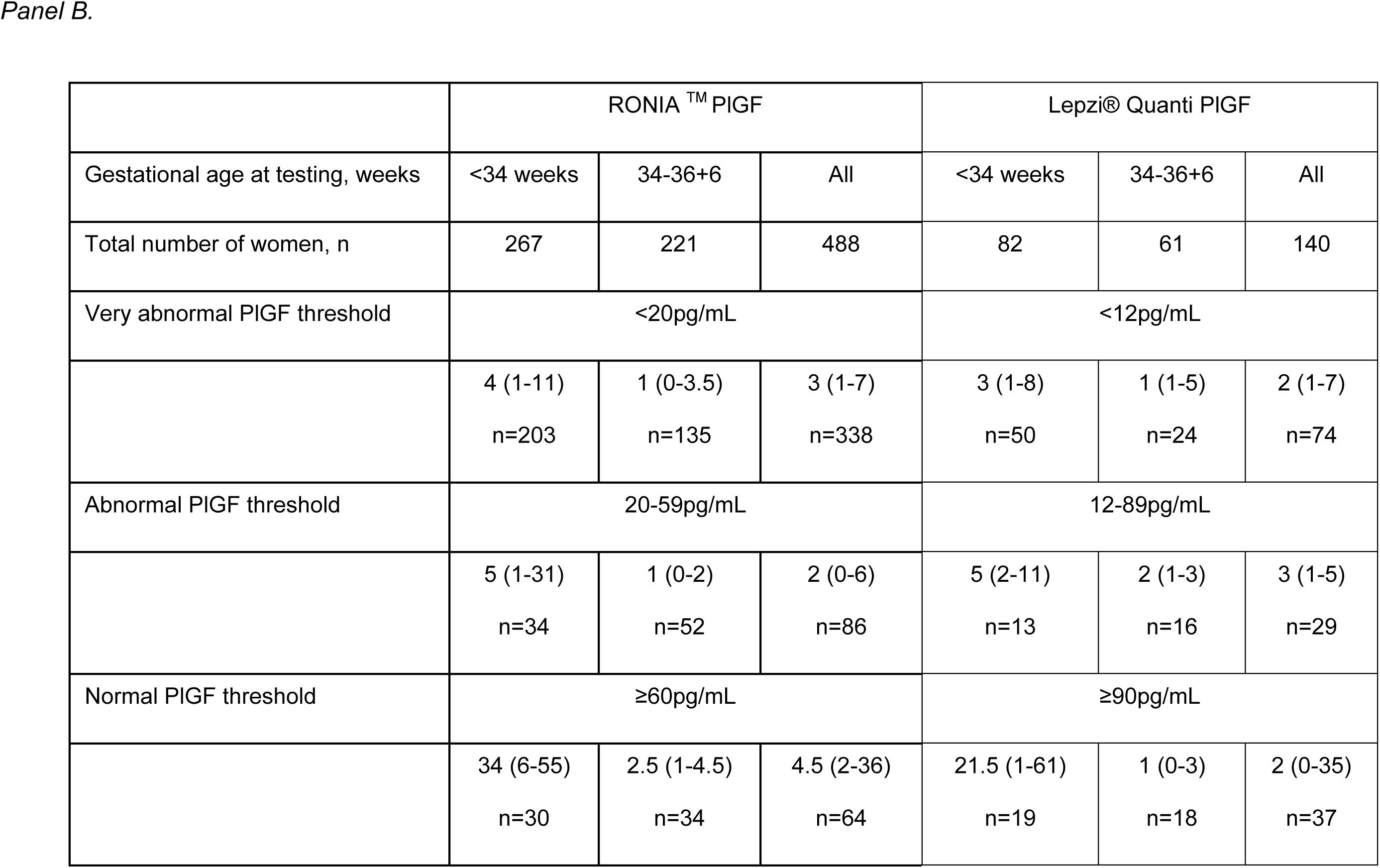
Time to delivery (median, IQR) stratified by RONIA^TM^ and Lepzi® Quanti PlGF concentration for all participants.

## Discussion

Whole blood RONIA^TM^ and Lepzi® Quanti PlGF both accurately identified women admitted with hypertension and/or clinical features of pre-eclampsia who were unlikely to have a serious adverse event, especially in those tested <34 weeks’ gestation, in a challenging LMIC environment. For maternal and perinatal composite outcomes this was represented by sensitivities of 94.9% and 100.0% respectively (RONIA^TM^), and 100.0% for both (Lepzi® Quanti). Median time to delivery was markedly shorter for women tested <34 weeks with abnormal RONIA^TM^ and Lepzi® Quanti PlGF values (compared to those with normal results).

Mortality and morbidity in women admitted in Sierra Leone is extremely high: in this study there were 17 (3.5%) maternal deaths, 50 eclampsia cases and 106 perinatal deaths. These are the largest numbers reported in any study investigating PlGF to date, and confirms that both RONIA ^TM^ and Lepzi® Quanti PlGF identify adverse events. Rule-out characteristics for severe events were greatest for women presenting at <34 weeks’ gestation but remained reliable, and could benefit those presenting up to 37 weeks’ gestation, enabling stratified surveillance and timing of delivery. However, most women with a very abnormal PlGF did not die, which explains the low specificities reported for both tests.

A strength of this study is conducting it in a setting with a high prevalence of maternal and perinatal adverse outcomes, compared to previous studies which have informed UK national guidelines(4, 5, 12), where no cases of maternal mortality and few stillbirths have occurred. By contrast, the current study enables confirmation that POC-PlGF can predict most of these events. The only case where a woman died with a normal RONIA^TM^ PlGF value was due to another cause, and no maternal deaths occurred with a normal Lepzi® Quanti PlGF.

This first study to evaluate whole blood POC-PlGF testing demonstrated that it is feasible and accurate, in a hospital in Sierra Leone, a country with high reported maternal (443/100 000(13)) and neonatal (3/1000(14)) mortality rates, challenging infrastructure (38% of health facilities lack reliable electricity access(15), required for device charging and test cassette refrigeration) and was acceptable to women (only four women refused). As diagnostic criteria for pre-eclampsia requires resources that were not always available (e.g. dipstick for proteinuria and ultrasound scans for growth) we were unable to stratify analysis by suspected and confirmed pre-eclampsia although the maximal mean BP was extremely high (173/118mmHg). PlGF concentrations were not revealed to study participants’ clinical teams so that test results could not influence provision of routine care, including decisions for delivery.

Accurate estimation of gestational age was challenging given limited access to ultrasound scans, and delayed presentation for antenatal care. Determination of gestational age is also more difficult in pre-eclampsia with growth restricted babies, so biomarker-assisted diagnosis to target delivery adds value; it is justified to deliver earlier gestations with very abnormal PlGF, whilst avoiding unnecessary early delivery in those with normal PlGF. Despite challenges with follow-up we were able to obtain data in over 90% of participants.

PlGF test performance has been demonstrated prospectively in LMIC settings; a study in Mozambique confirmed that abnormal compared to normal PlGF in suspected pre-eclampsia was associated with a significantly higher risk of pre-eclampsia diagnosis, shorter time-to-delivery and perinatal death(8). In a subsequent pilot study, one-third of women with very abnormal PlGF had a stillbirth; AUC for stillbirth was 0.78 (95%CI 0.70-0.86), with a sensitivity of 76.9% (95%CI 60.7 – 88.9%) and specificity 74.1% (95%CI 68.0 – 79.5%)(9). In our study, where twice the number of stillbirths occurred, both RONIA^TM^ and Lepzi®® Quanti PlGF demonstrated superior sensitivity of 98.8%, and 95.8% respectively, although more modest specificity, 19.8% and 31.5% respectively. In India, a small prospective study (n=50) found that abnormal SFlt-1/PlGF results were significantly associated with pre-eclampsia with severe features (90.91% vs 8.00%, p < 0.0001) and a higher composite maternal complication rate (18.18% vs 0%, p = 0.04), compared to 40.0% vs 8.0% in women tested <34 weeks’ gestation in our study (10).

All studies to date have used benchtop analyzers for PlGF-based testing, requiring pre-analysis centrifugation, challenging in Sierra Leone, and other low-resource settings. PlGF alone and SFlt-1/PlGF ratio are both recommended by NICE, with no differences in their ability to predict outcomes(16, 17) However, POC testing would be significantly more difficult with two assays and is not currently available. In this study, we have prospectively demonstrated accurate rule-out of adverse maternal and neonatal events, based on whole blood samples, demonstrating test accuracy and feasibility of both RONIA ^TM^ and Lepzi® Quanti PlGF in a setting where there is huge potential for impact.

### Perspectives: Implications for clinicians and future research

This is the first prospective observational study of novel whole blood POC-PlGF testing, which could enable individualized risk stratification, targeted allocation of scare resources, and planned early delivery from 34 weeks, known to reduce stillbirth(18). Conversely, high sensitivity and NPV, facilitates rule-out of adverse maternal and neonatal outcomes, providing justification to manage women expectantly in settings with limited neonatal care. POC testing is likely to be considerably cheaper than currently recommend PlGF-based tests, and companies are targeting LMIC settings. Given the expense of current devices and tests (>$2200/ device; >$30 USD/ test) (8), it is likely that these POC tests could be adopted worldwide. Whole blood POC-PlGF testing should be implemented and evaluated as part of pre-eclampsia care pathways alongside other evidence-based strategies of early detection and delivery.

## Conclusion

This study indicates that both whole blood RONIA^TM^ and Lepzi® Quanti PlGF testing could enable appropriate management and surveillance to be targeted to those at greatest risk, whilst avoiding unnecessary, costly intervention with a normal result, and is feasible in challenging settings.

## Data Availability

The dataset will be available to appropriate academic parties on request to the chief investigator (AHS) in accordance with the data sharing policies of King's College London.

## Acknowledgements

All authors contributed to the conception and planning of the work. KK, RC, MM contributed to carrying out the work. KK led the writing. All authors contributed to analysis and interpretation of the work, writing the manuscript, final approval of this version, and agree to be accountable for all aspects of the work.

## Supplemental Materials

### Supplemental Figures

Figure S1 STARD Checklist.

Figure S2 Hypertension category and adverse maternal and perinatal events stratified by PlGF category in women tested 34-36+6 weeks’ gestation.

### Supplemental Tables

Table S1 Details of adverse maternal outcomes, based on miniPIERS consensus

Table S2 Details of adverse perinatal outcomes

Table S3 RONIA^TM^ and Lepzi® Quanti PlGF prediction of maternal death (individual component of primary maternal composite outcome) based on PlGF thresholds of <20pg/mL and <60pg/mL, and <12pg/mL and <90pg/mL respectively

Table S4 RONIA^TM^ and Lepzi® Quanti PlGF prediction of eclampsia (individual component of primary maternal composite outcome) based on PlGF thresholds of <20pg/mL and <60pg/mL, and <12pg/mL and <90pg/mL respectively.

Table S5 RONIA^TM^ and Lepzi® Quanti PlGF prediction of stillbirth (individual component of perinatal composite outcome) based on PlGF thresholds of <20pg/mL and <60pg/mL, and <12pg/mL and <90pg/mL respectively.

Table S6 RONIA^TM^ and Lepzi® Quanti PlGF prediction of neonatal death before discharge (individual component of perinatal composite outcome) based on PlGF thresholds of <20pg/mL and <60pg/mL, and <12pg/mL and <90pg/mL respectively.

Table S7 RONIA^TM^ and Lepzi® Quanti PlGF prediction of small-for-gestational age (SGA) <10th centile based on PlGF thresholds of <20pg/mL and <60pg/mL, and <12pg/mL and <90pg/mL respectively. SGA is calculated for live births only.

Table S8 RONIA^TM^ and Lepzi® Quanti PlGF prediction of small-for-gestational age (SGA) <3rd centile based on PlGF thresholds of <20pg/mL and <60pg/mL, and <12pg/mL and <90pg/mL respectively.

Table S9 RONIA^TM^ and Lepzi® Quanti PlGF prediction of delivery within 7 days of test based on PlGF thresholds of <20pg/mL and <60pg/mL, and <12pg/mL and <90pg/mL respectively.

Table S10 RONIA^TM^ and Lepzi® Quanti PlGF prediction of delivery within 14 days of test based on PlGF thresholds of <20pg/mL and <60pg/mL, and <12pg/mL and <90pg/mL respectively.

Table S11 RONIA^TM^ and Lepzi® Quanti PlGF prediction of delivery less than 37 weeks’ gestation based on PlGF thresholds of <20pg/mL and <60pg/mL, and <12pg/mL and <90pg/mL respectively.

Table S12 RONIA^TM^ and Lepzi® Quanti PlGF prediction of composite maternal outcome, excluding twin pregnancies, based on PlGF thresholds of <20pg/mL and <60pg/mL, and <12 and <90pg/mL respectively.

Table S13 RONIA^TM^ and Lepzi® Quanti PlGF prediction of composite perinatal outcome, excluding twin pregnancies, based on PlGF thresholds of <20pg/mL and <60pg/mL, and <12 and <90pg/mL respectively.

## Novelty and relevance

### What is new?

For the first time, we have prospectively demonstrated that two novel whole blood point-of-care PlGF tests can accurately rule out maternal death, eclampsia and stillbirth in women with suspected pre-eclampsia in a low-resource setting.

### What is relevant?

Pre-eclampsia accounts for 25% of maternal death in LMICs, and is linked to a 2-6 fold increased risk of hypertension and cardiovascular disease. Most deaths are preventable with early detection and risk stratification, to enable timely management.

### Clinical/ Pathophysiological implications

Point-of-care PlGF can accurately rule-out serious pre-eclampsia-related adverse pregnancy outcomes, and could support clinicians to plan timing of delivery, especially valuable in settings with limited neonatal resources.

